# Characterisation of 3000 patient reported outcomes with predictive machine learning to develop a scientific platform to study fatigue in Inflammatory Bowel Disease

**DOI:** 10.1101/2025.01.18.25320777

**Authors:** Rebecca Hall, Robert J Whelan, Peter D Cartlidge, Emily F Brownson, Craig Mowat, John P Seenan, Jonathan C MacDonald, Iona AM Campbell, Cher S Chuah, Gwo-Tzer Ho

## Abstract

**Background:** Fatigue is commonly identified by IBD patients as major issue that affects their wellbeing. This presentation, however, is complex, multifactorial and mired in clinical heterogeneity.

**Aims/Methods:** We prospectively captured patient reported outcomes (PROs) from 2 current IBD biomarker studies in Scotland with ∼100 clinical metadata points; and an international dataset (that includes non-IBD healthy controls) using CUCQ32, a validated IBD questionnaire to generate a contemporaneous dataset of fatigue and overall wellbeing (2021-2024) and utilized 6 different machine learning (ML) approaches to predict IBD-associated fatigue and patterns that may aid future stratification to human mechanistic and clinical studies.

**Results:** In 2 970 responses from 2 290 participants, CUCQ32 were higher in active IBD vs. remission; and in remission, higher than in non-IBD controls (both p<0.0001). CUCQ32-specific fatigue score significantly correlated to all CUCQ32 components (p=2.9 x 10^-28^ to 3.2 x 10^-147^). During active IBD, patients had significantly more fatigue days compared to those in remission and non-IBD controls (medians 14 vs. 7 vs. 4 [out of 14 days]; both p<0.0001). We determine a threshold of ≥10/14 days of fatigue as clinically relevant - Fatigue_high_. Overall, 72.8% (863/1185), 45.0% (408/906) and 13.7% (46/355) responses in active, remission and non-IBD controls were in Fatigue_high_. Using train-validate-test steps, we incorporated all available metadata to generate ML-models to predict Fatigue_high_. The 6 ML models performed similarly (all 6 models AUC of ∼0.70). SHapley Additive exPlanations (SHAP) analysis revealed that each algorithm places different importance on variables with seasonality, biologic drug levels, BMI and gender identified as factors. ML prediction of Fatigue_high_ in patients in biochemical remission (CRP<5 mg/l and calprotectin <250μg/g) was more challenging with AUC of 0.66-0.61.

**Conclusion:** We provide a comprehensive patient involvement-ML-pathway to predict IBD-associated fatigue. Our data suggests a large ‘hidden’ pathobiological component and current work is in progress to integrate deep molecular data and build a clinical-scientific ML model as a step towards better understanding of IBD-associated fatigue.

## Introduction

Wellbeing is complex and multifaceted, encompassing many dimensions of quality of life, physical and mental health. Improving wellbeing is an important patient goal and a treatment target in IBD^1^. Recently, patient reported outcomes (PROs) as better reflection of wellbeing, are now mandated by drug regulatory bodies such as FDA/EMA as necessary endpoints in new clinical trials ^2, 3^.There are many increasingly potent anti-inflammatory therapies with clear evidence in their ability to improve and treat gut inflammation. However, the extent that they improve wellbeing remains in question. Fatigue is a key case in point.

In IBD, fatigue is often reported as the most troublesome and widely reported symptom^4^. The patient-focused initiative James Lind Alliance review places fatigue as a top priority for IBD research^5^. The burden of fatigue is notable with a prevalence of 50% in IBD patients^6^. Despite objective improvements in gut inflammation in response to drug treatments, many patients continue to experience significant levels of fatigue^7^. Pertinently, there are only a few interventional studies that scientifically target IBD-associated fatigue, with poor results^8–10^ or are small and open-labelled in nature^11^. The scientific basis of IBD-associated fatigue remains poorly understood and is a major unmet need in IBD.

In this study, we aim to capture real-world PROs incorporating social, psychological and emotional health in IBD and non-IBD subjects and investigate how they relate to IBD-associated fatigue. We use the patient-reported Crohn’s and Ulcerative Colitis Questionnaire-32 (CUCQ32) questionnaire that is relevant to both Crohn’s disease (CD) and Ulcerative colitis (UC), as the initial basis to capture wellbeing and link this data with multiple levels of clinical and laboratory data in IBD. We harmonised CUCQ32 data from 2 sources: with consented data from our ongoing studies in our IBD units in Scotland; and a larger online cohort across UK and internationally. We then reduce the dimensionality of the complex PROs of wellbeing and clinical IBD activity with IBD-associated fatigue in this dataset and employed machine learning (ML) algorithms incorporating our captured datapoints to predict fatigue using a simple patient-reported threshold of fatigue.

Given the complexities of fatigue, we aim to develop a ML-assisted model that incorporates expansive features including seasonality, different drug therapies and body mass index for examples. We then envisage that with this initial approach, we can build towards a platform where we can identify deeper clinical patterns and factors; groups of patients that we can further incorporate molecular and biological data to explain the inherent variability of fatigue and finally, to use this as a platform to develop targeted human experimental approaches to investigate the pathobiology of IBD-associated fatigue.

## Results

### Prospective CUCQ32 data capture in 2 IBD patient cohorts (2020-24) with aligned clinical metadata – Cohorts 1 and 2

CUCQ32 captures 4 domains of health pertaining to gastrointestinal symptoms, psychological, social and general well-being (Table 1, Appendix 1) with a score range from 0-272 (higher score = worse quality of life). We recorded CUCQ32 PROs in the MUSIC study (‘Mitochondrial DAMPs as mechanistic biomarkers in IBD; www.musicstudy.uk) and the GI-DAMPs (Investigation into Gastrointestinal Damage-Associated Patterns in IBD) (from hereon, Cohorts 1 and 2) (Figure 1a, b).

**Figure 1.**
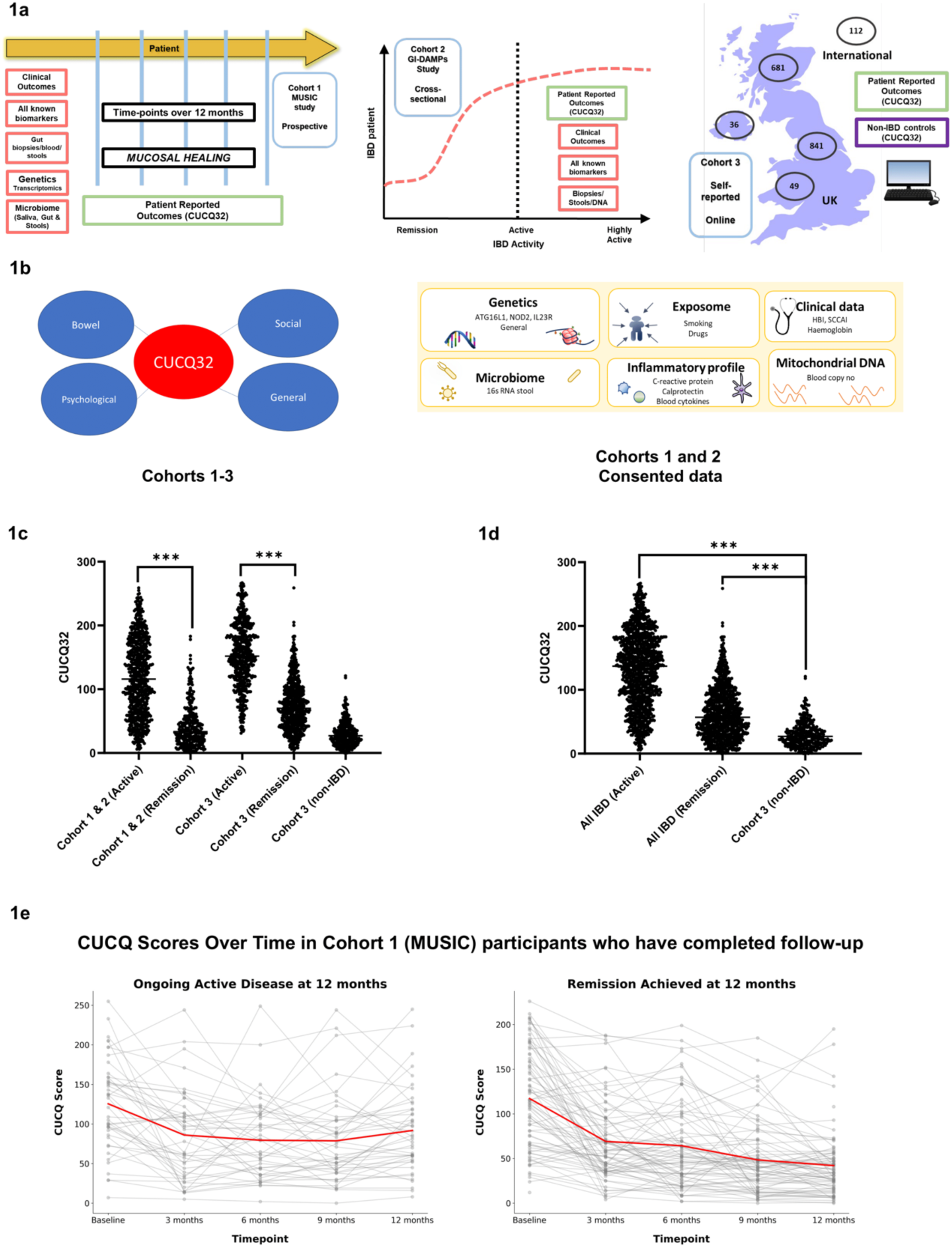
Overview of the study cohorts 1a: Overview of the 3 study cohorts - left: MUSIC cohort (Cohort 1) middle: GI-DAMPs cohort (Cohort 2) right: Online survey cohort (Cohort 3). 1b:CUCQ-32 questionnaire dimensions; Overview of data collection in Cohorts 1 & 2. 1c: CUCQ-32 scores are higher in patients with active I8D. 1d: CUCQ-32 scores are higher in patients with IBD compared to non-I8D controls, even in disease remission. 1e: Evolution of CUCQ-32 scores over time in patients who have ongoing disease activity vs. remission in the MUSIC cohort. Red line representing the mean. (Cohort 3)

**Table 1:**
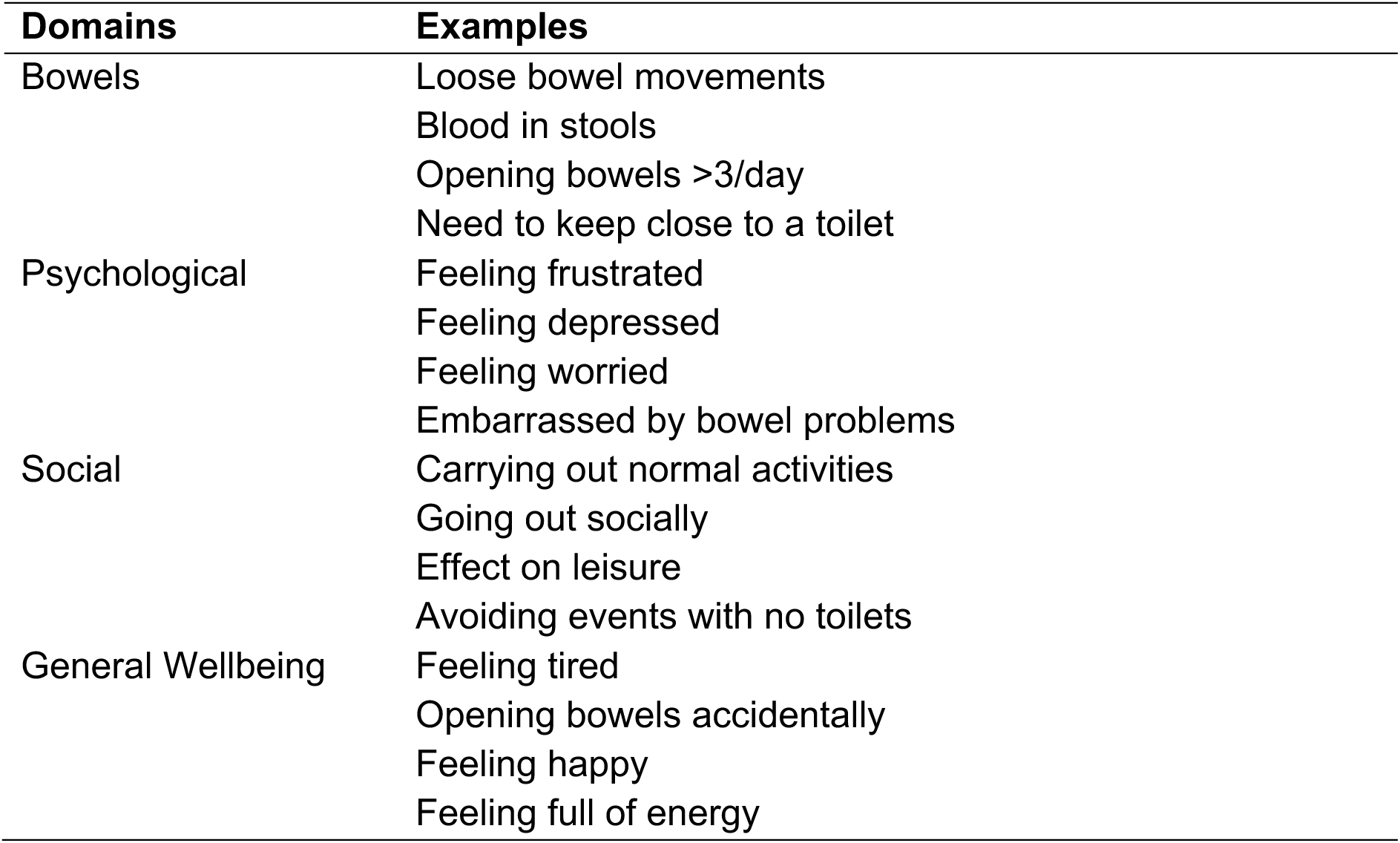
The CUCQ32 scoring 0-272 encompasses 4 domains^25^.

MUSIC is a prospective cohort study that incorporates 5 clinical timepoints (every 3 months) over a 12-month follow-up period. At the entry point to MUSIC, all patients have documented active IBD, confirmed endoscopically and with accompanying imaging and positive stool calprotectin tests. GI-DAMPs is a cross-sectional study where IBD patients with active disease or in remission are recruited. Both are scientific studies where IBD patients are recruited from Glasgow, Edinburgh and Dundee in Scotland, United Kingdom (2020-present).

Hence, both MUSIC and GI-DAMP studies provide a diverse landscape of IBD presentations over time with prospectively recorded patient-reported outcomes in CUCQ32. In these consented cohorts, ∼100 lines of metadata are available including clinical (disease activity, extent, severity, treatment), laboratory (e.g. full blood count, C-reactive protein, calprotectin), body mass index, exposome (e.g. smoking), drug therapy at sampling and seasonality. These are paired to CUCQ32 data entry time points (Appendix 2, 3). In these studies, we also have biological data such as genetics and stool microbiome in subsets of patients (Figure 1b). In the MUSIC data, prospective endoscopic mucosal disease activity (SES-CD and UCEIS/Endoscopic Mayo Score are available for CD and UC respectively) are available that will allow us to assess endoscopic mucosal healing in relation to CUCQ32. The demographics and cohort characteristics are shown in Table 2.

**Table 2:**
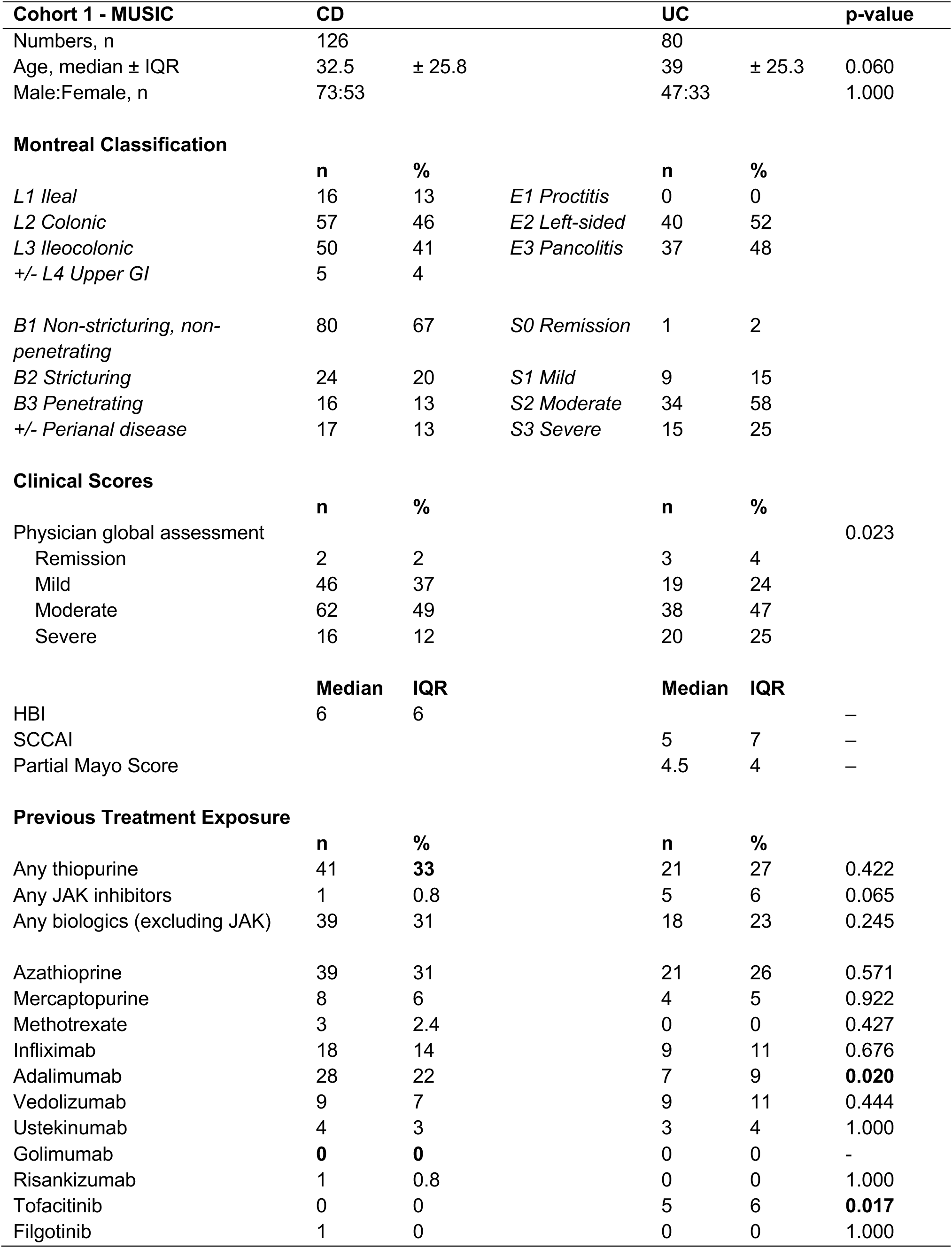
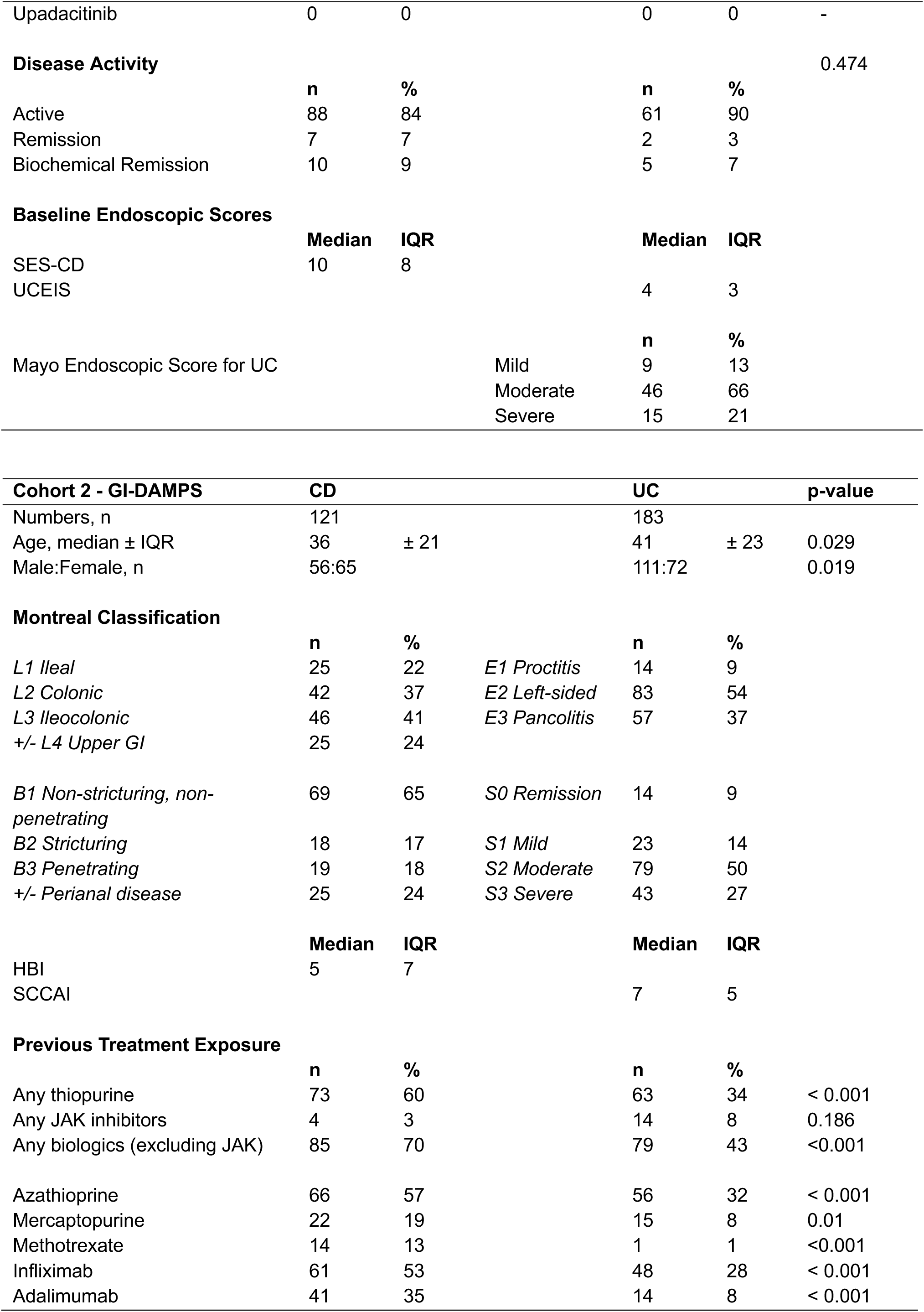
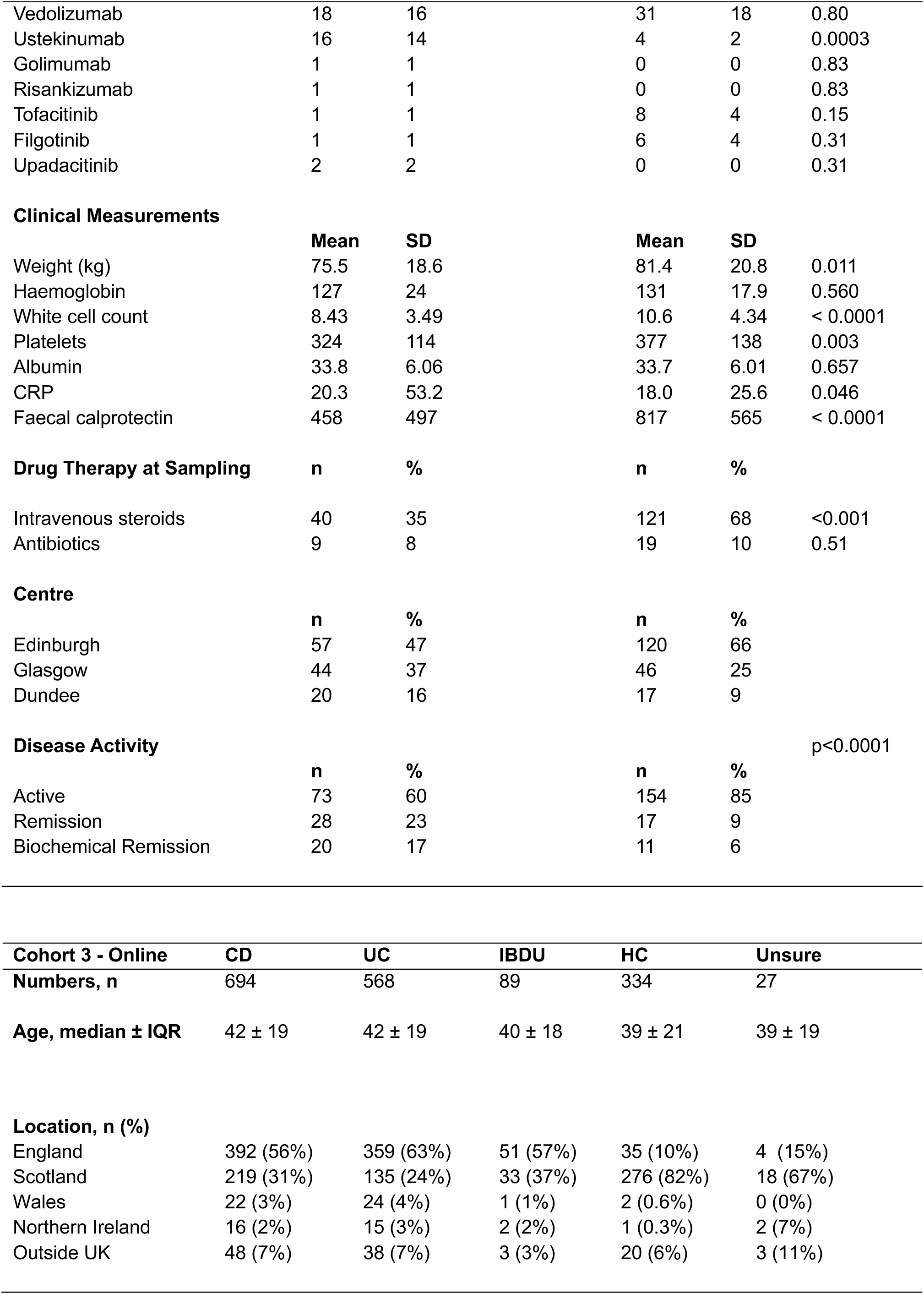
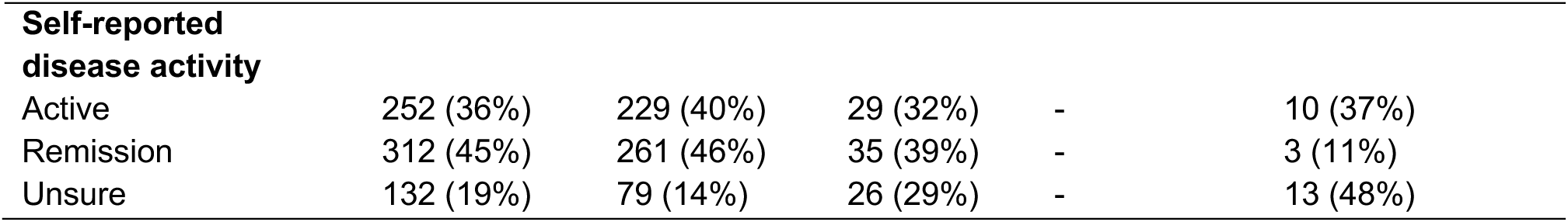
Demographics and clinical characteristics of Cohorts 1 (MUSIC), 2 (GI-DAMPs) and 3 (self-reported online).

### CUCQ32 in self-reported IBD and non-IBD patient cohorts (2023-24) – Cohort 3

In addition to this, we adopted an online self-reported approach (accessible via https://forms.office.com/e/aJWUEC4UUN) for IBD patients in the UK and internationally (Cohort 3) (Figure 1a). Here, we used our IBD networks to invite patients to provide CUCQ32 scores and how they relate to their disease activity. Our network included patient charitable groups such as Crohn’s Colitis UK, Catherine McEwan Foundation and Cure Crohn’s Colitis; via our Edinburgh IBD Science network (@Edin_IBDScience) and the University of Edinburgh; directly in our IBD clinics in Glasgow, Edinburgh and Dundee; and finally via our Patient-Public Involvement groups.

To retain anonymity, we avoided patient-identifiable information and recorded minimal demographic and disease-specific data. Participants were given the opportunity to provide further any further relevant information and to highlight patient-focused areas of importance in an anonymous capacity. Using this approach, we included 1721 self-reported CUCQ32 responses from a total of 1358 IBD patients (697, 571 and 90 CD, UC and IBD-U respectively; as of April 2024). We captured data from the UK (681, 841, 49 and 36 from Scotland, England, Wales and Northern Ireland respectively) and internationally (112 from Outside UK). To provide a non-IBD benchmark, we also collected CUCQ32 data from self-nominated non-IBD controls (‘*I don’t have IBD – I am a healthy control’*) and those with bowel symptoms but do not have a formal diagnosis of IBD; 336 and 27 responses respectively.

### IBD well-being as defined by CUC3Q32 in active and remission states

In total, we studied a cohort of 2 290 subjects with 2 970 CUCQ32 data. We partitioned our data by clinical disease activity status (active vs. remission vs. biochemical remission) in Cohorts 1 and 2. Active vs remission was defined as the presence of IBD symptoms, recorded by the clinician at the study visit. Biochemical remission was defined as being in remission and having a stool calprotectin of <250ug/g and a CRP of <5 mg/L. Here, median CUCQ32 scores were significantly higher in patients with active disease (133 vs 40 vs 39 for active, remission and biochemical remission; analysis of variance F = 296, p<0.001).

Using a similar approach in Cohort 3, we also found significantly higher CUCQ32 in active disease (151 vs. 70 in active vs remission; p<0.001) (Figure 1c). As there is no ‘normal’ for CUCQ32, we combined all our cohorts to provide a descriptive spread of our data and to benchmark our CUCQ32 data in IBD against a group of individuals with no IBD and self-reported to be ‘healthy’ in Cohort 3 (All IBD – n=1151 and 1061 responses in active and remission respectively). Here, we calculated the coefficient of variation for CUCQ32 which can be quantified by the ratio of the standard deviation to the mean. The inter-patient variation of IBD and non-IBD healthy controls ranged from 41% to 67%.

It is noteworthy that median CUCQ32 in IBD patients in remission is significantly higher than in non-IBD controls, 56 vs. 27 (ANOVA with post-hoc Bonferroni correction, p<0.001) (Figure 1d). For Cohort 1, we prospectively tracked 147 IBD patients with completed 12-month follow-up with 5 serial CUCQ32 data. Here, there are notable inter-patient variations of CUCQ32; and their respective trajectories although IBD patients that achieve clinical remission at end of 12 months follow-up demonstrate a downward trend of CUCQ32 (Figure 1e).

### CUCQ32 is significantly correlated with clinical activity indices for UC and CD

In Cohorts 1 and 2, CUCQ32 were significantly correlated with clinician-based assessment of IBD activity of the patient; Harvey-Bradshaw Index (HBI) and Simple Clinical Colitis Activity Index (SCCAI) for CD and UC respectively (r=0.76 and 0.82; both p<0.001 respectively) (Figures 2a and b). In these cohorts, CUCQ32 were also significantly higher in IBD patients with high calprotectin (>250 ug/g) and C-reactive protein (>10 mg/L) (Figures 2c and d). In Cohort 1 (MUSIC study) patients who have completed 12 months of follow-up, there was no difference in CUCQ32 between the groups of IBD patients that achieved endoscopic mucosal healing and those who did not (median CUCQ score of 68, 62 in complete mucosal healing versus no mucosal healing respectively, Figure 2e).

**Figure 2.**
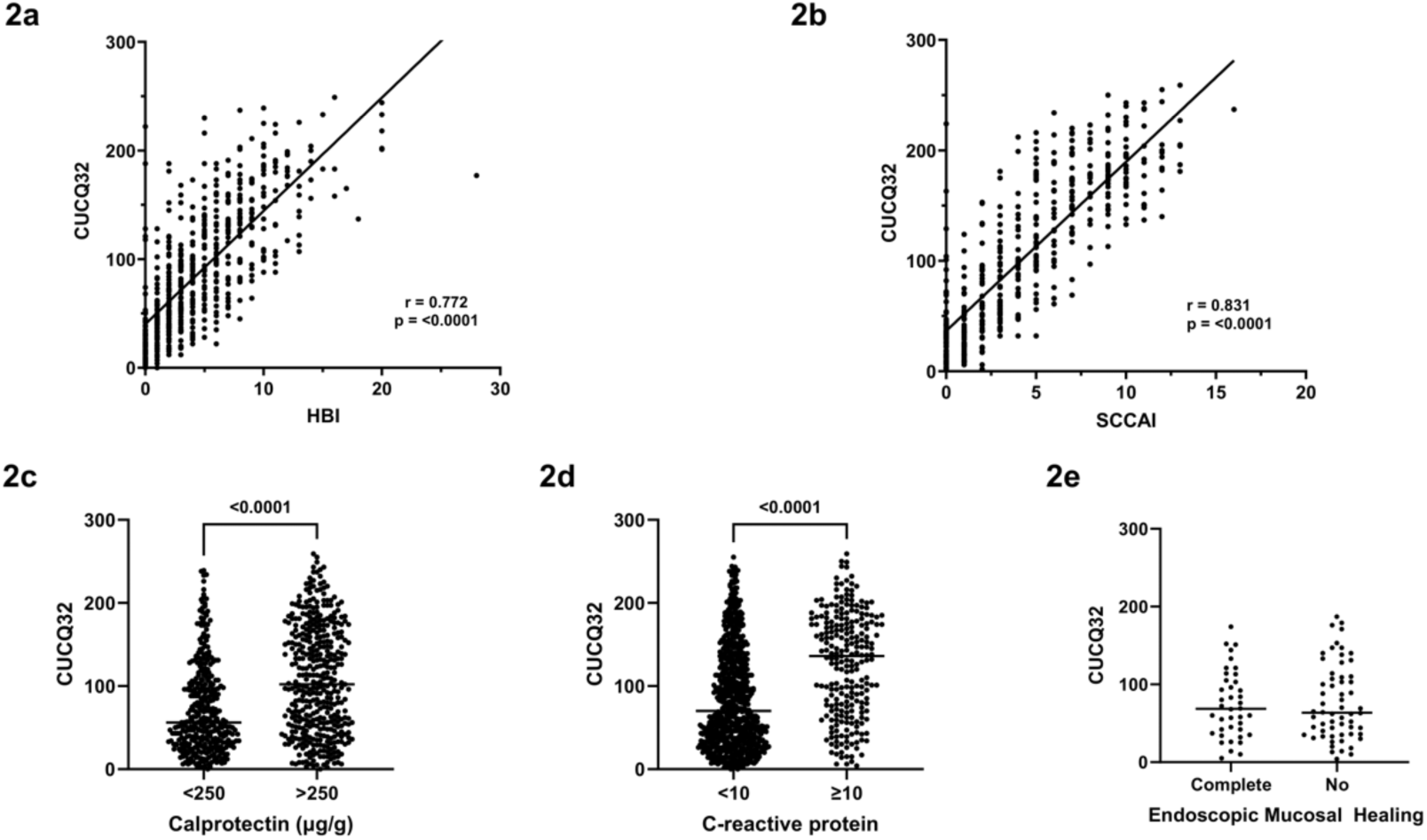
CUCQ 32 scores and correlation with clinical, laboratory and endoscopic parameters. 2a: CUCQ-32 scores are highly correlated with Harvey-Bradshaw Index scores. (Crohn’s disease) 2b: CUCQ-32 scores are highly correlated with Simple Clinical Colitis Activity Index scores. (Ulcerative colitis) 2c: CUCQ-32 scores are significantly higher in patients with raised faecal calprotectin values >250 ug/g 2d: CUCQ-32 scores are significantly higher in patients with elevated C-reactive protein values >10mg/L. 2e: No significant difference was seen in CUCQ-32 scores in patients who achieved complete mucosal healing vs patients without, in Cohort 3 (MUSIC).

Collectively, our data shows that patient-reported outcomes using CUCQ32 are good indicators of clinician-based assessment and typical clinical measurements of IBD activity. Our CUCQ32 data collected within a consented clinical study framework agreed broadly with the wider self-reported data on disease activity. Of interest, IBD patients who are in clinical remission have higher CUCQ32 scores (worse quality of life) when benchmarked to individuals without IBD (Figure 3a).

**Figure 3.**
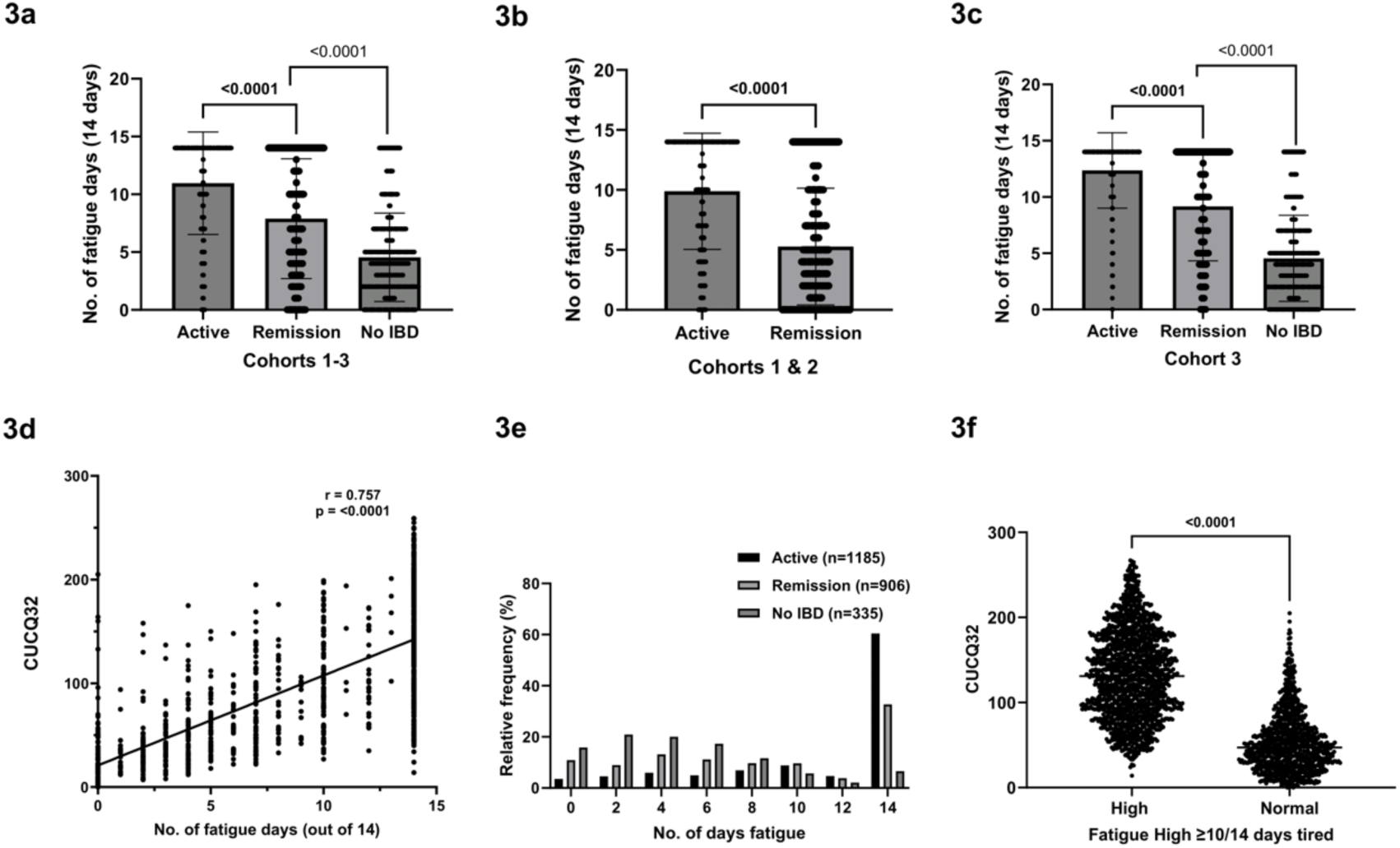
3a: Median number of fatigue days were significantly different between patients with active IBD, patients in remission and non-lBD controls (All cohorts combined) 3b: Median number of fatigue days were significantly higher in patients with active disease. (Cohorts 1 & 2). 3c: Median number of fatigue days were significantly different between patients with active IBD, patients in remission and non-lBD controls. (Cohort 3 only). 3d: Number of fatigue days reported in CUCQ question 5 was significantly correlated with overall CUCQ-32 scores. 3e: Distribution of reported number of days with fatigue demonstrating non-normal distribution. Median number of days was 10 and used to convert this continuous variable into binary classification of Fatigue-high and Fatigue-low. 3f: Overall CUCQ-32 scores are significantly higher after conversion into binary classification.

### Fatigue is significantly correlated to all CUCQ32 dimensions of wellbeing

Within the CUCQ32, we focused on the specific area of fatigue where our subjects were asked ‘how many days in the last 2 weeks did you feel tired?’ with a score of 0-14 days (Question 5; Appendix 1). This data is significantly correlated to all the 32 questions in CUCQ32 in our dataset (P_minimum_ 2.97 x 10^-28^ to P_maximum_ 3.18 x 10^-147^) with the strongest correlation observed with Question 2, ‘On how many days over the last two weeks have you felt generally unwell?’ (Appendix 3).

In our dataset, the top 3 most significantly correlated questions are: (1) ‘On how many days have you felt generally unwell?’ (Question 2), (2) ‘On how many days over the last two weeks have you felt full of energy?’ (Question 16); and (3) ‘On how many nights have you been unable to sleep well?’ (Question 12). This data underscores the central importance of fatigue in overall quality of life in IBD and the potential to reduce the dimensionality of the CUCQ32 dataset for further analysis on fatigue.

In Cohort 3, where participants were asked to list areas of importance where there is greatest unmet need, 1 036 responses were recorded. Fatigue (25.1%) was ranked highest, followed by stress (19.2%) and diet/alcohol (14.3%). This finding is in close agreement with a parallel UK-wide exercise conducted by Crohn’s and Colitis UK in 2024 (https://crohnsandcolitis.org.uk/media/ozvb3rch/top-10-impacts-of-living-with-crohns-and-colitis.pdf).

### IBD patients have more fatigue days in active and remission states compared to non-IBD controls

Firstly, we performed our analysis in all 3 cohorts combined and found that IBD patients with active IBD had significantly more fatigued days compared to those in remission (medians 14 vs. 7 days in 1151 and 1061 responses respectively, p<0.0001) (Figure 3a, b). Of interest, IBD patients in remission also had significantly higher fatigue days compared to non-IBD control group (medians of 7 vs. 4 days in 1061 and 339 responses respectively; p<0.0001). These findings are maintained when Cohorts 1 and 2, and Cohort 3 (online) were considered separately (Figures 3b, c). Fatigue days are highly correlated to CUCQ32 (r=0.73, 2,902 paired data points, p<0.0001) (Figure 3d). In summary, our IBD data is skewed towards fatigue with medians of 14, 7 and 4 days (IQR 8-14, 3-7 and 2-7 days) in active, remission and non-IBD controls respectively (Figure 3e).

### Defining the clinical threshold of IBD-associated fatigue

Given the non-normal distribution of reported fatigued (Figure 3e), we determine a threshold of ≥10/14 days of fatigue using the median value (median 10 days; mean 9 days for comparative reference) – from here on Fatigue_high_. In our combined datasets (Cohorts 1, 2 and 3), we found an overall prevalence for Fatigue_high_ of 53.9% (1 602 out of 2 970). Here, we found 75.2% (866/1151), 43.6% (463/1061) and 14.2% (48/339) responses in active, remission and non-IBD controls respectively to fall in the Fatigue_high_ (Figure 3e). In Fatigue_high_ group, CUCQ32 scores were significantly higher (130 vs. 38; p<0.0001 in 1377 and 1174 responses respectively) (Figure 3f).

### Machine-learning approach to predict IBD-associated fatigue

Using all our available clinical data in Cohorts 1 and 2 with total (including CUCQ32-patient reported outcomes, seasonality, BMI, drug treatments, disease phenotype and all hospital-based blood/stool tests, full list in Appendix 4) and 1215 data points, we employed six different machine learning approaches (XGBoost, random forest, AdaBoost, multilayer perceptron classifier, support vector classifier, logistic regression) to develop models to predict Fatigue_high_ in IBD. We also further investigated the prediction of fatigue in IBD patients in biochemical remission (Figures 4a).

**Figure 4.**
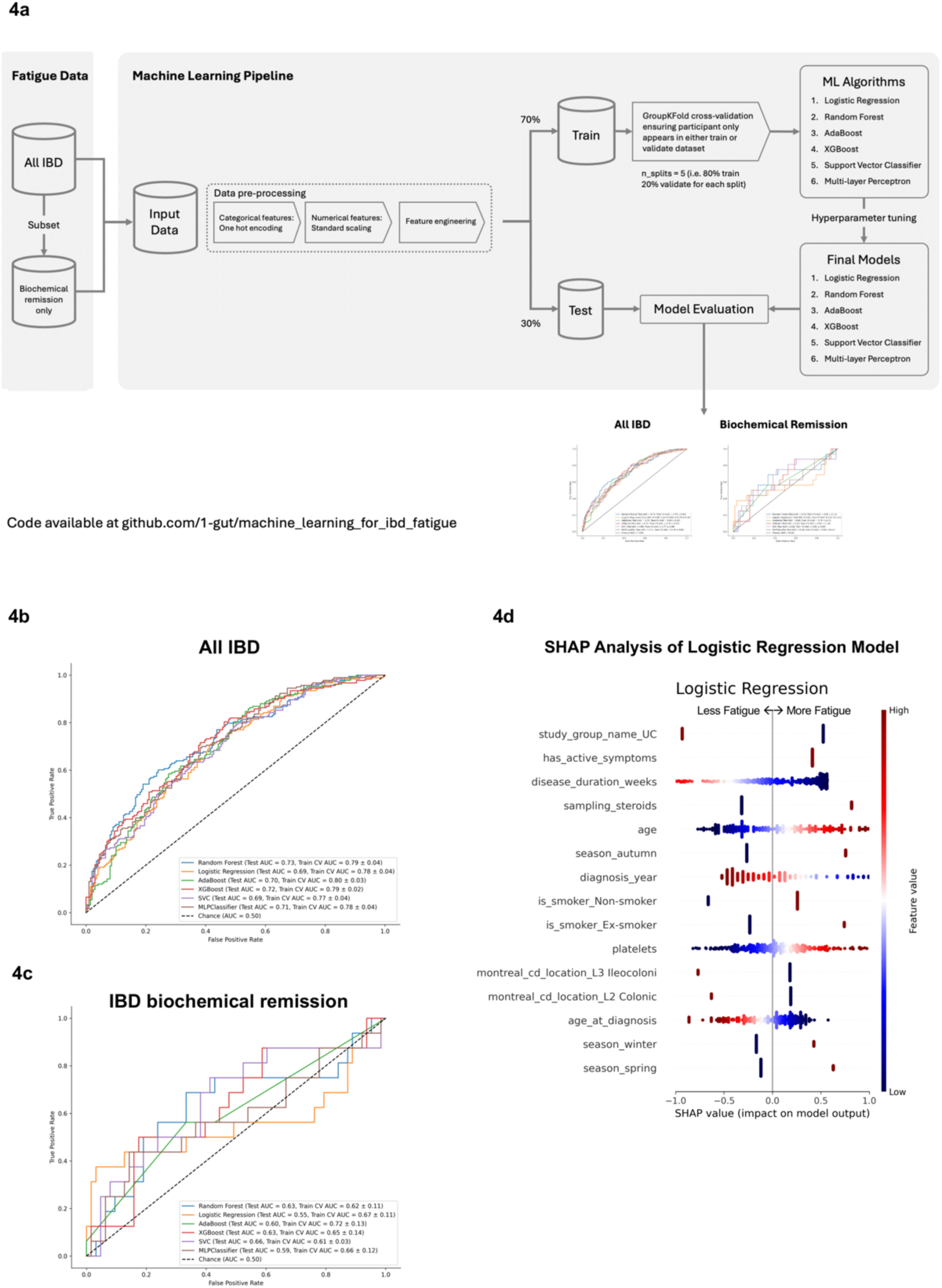
4a: Summary of machine learning pipeline. 2 pipelines were run - one for all lBD patients in Cohort 1-2 and the second for those identified in biochemical remission, defined as the absence of active IBD symptoms and calprotectin <250ug/g and CRP <5 mg/dl. GroupKFold cross-validation was used in model construction. 4b: Summary of model performance in the all lBD cohort using the metric of area under the receiver operating characteristic curve (AUC). All models tested demonstrated similar performance. 4c: Summary of model performance in the biochemical remission cohort using the metric of area under the receiver operating characteristic curve (AUC). These models performed worse compared to the all IBD cohort. 4d: Shapley additive explanation values for the logistic regression demonstrating the modelling of seasonality. For example, season_autumn (which either has values of O or 1) - a value of 1 (red) indicates that the model is more likely to predict fatigue.

We employed train-validate-test steps with GroupKFold cross-validation and showed that these models performed similarly with areas under the curve (AUC) ranging from 0.69 to 0.73; maximum AUC of 0.73 was achieved in the Random Forest model (Figure 4b) (Table 3). ML prediction of IBD-fatigue_high_ in patients in biochemical remission (CRP<5 mg/l and calprotectin <250μg/g) was more challenging with AUC of 0.66-0.61 (Figure 4c).

**Table 3:** Model performance of the 6 different machine learning approaches.

| **Model** | **Recall**<br>**(Sensitivity)** | **Precision**<br>**(Specificity)** | **Accuracy** | **AUC** |
| --- | --- | --- | --- | --- |
| Random Forest | 0.66 | 0.64 | 0.65 | 0.73 |
| XGBoost | 0.69 | 0.62 | 0.66 | 0.72 |
| MLPClassifier | 0.63 | 0.65 | 0.64 | 0.71 |
| AdaBoost | 0.63 | 0.65 | 0.64 | 0.70 |
| Support Vector Classifier | 0.61 | 0.65 | 0.63 | 0.69 |
| Logistic Regression | 0.63 | 0.63 | 0.63 | 0.69 |

### Analysis of the prioritisation of clinical factors by machine learning models

SHAP (SHapley Additive exPlanations) values are a powerful tool for interpreting machine learning models. They provide a way to understand the contributions of each input feature to the model’s predictions using a game theory approach^12^. Of interest in our dataset, SHAP analysis revealed that each model that we employed, prioritizes different variables (Supplementary Figure 1b, 1c); and seasonality (summer vs. winter) was modelled best by logistic regression (Figure 4d). However, other notable features included therapeutic drug levels (anti-TNF), BMI and biological sex. SHAP analysis showed high impact of active symptoms, blood markers of disease activity and use of steroids in the predictive modelling of Random Forest for Fatigue_high_ (Supplementary Figure 1b).

To summarise, in the all IBD cohort, clinical factors which drove the prediction of fatigue was generally reflective of active disease (for example, low haemoglobin, high platelet count). However, of interest, these factors were not useful in our patients in the IBD cohort who were in biochemical remission, where all the ML models performed poorly (Supplementary Table 1c).

## Discussion

We present one of the largest combined pool of prospectively captured real world’ patient reported outcomes into wellbeing across 3 contemporaneous cohorts with 2 970 responses from 2 290 participants – in UK and internationally, including CUCQ32 data from non-IBD control population as a benchmark. There are a few notable findings. Firstly, patient reported outcomes (‘*what patients tells clinicians*’) are highly correlated to clinician-based assessment (‘*what clinicians think of patients*’) and objective markers of gut inflammation. This data suggests that patient reported outcomes should be routinely captured and incorporated into clinical management of IBD care and stimulate an appraisal of whether a more ‘patient-dominant’ model of care should be considered.

Secondly, fatigue is a major feature of poor wellbeing in IBD. Patient reported fatigue are highly correlated to all other dimensions of wellbeing in CUCQ32 and in parallel reporting, our participants presented fatigue as the most common issue with an unmet need. Although active disease is well-recognized as a key contributory factor to fatigue, we showed that 43.6% of IBD patients in remission report high levels of fatigue. In our MUSIC study, where we carried out prospective endoscopic evaluation of disease activity, 27% of patients have significant levels of fatigue even when complete mucosal healing was achieved. This observation is also present in many immune-mediated diseases (IMIDs) such as rheumatoid arthritis, sarcoidosis and systemic lupus erythematosus; and in long Covid19 pointing towards an immune-mechanisms beyond organ specific-involvement and inflammation^13^.

Why patients develop significant levels of fatigue are poorly understood and often regarded as multifactorial in nature. Here, we sought to develop a simplified approach that stratify fatigue in larger datasets. Within the framework of the CUCQ32, we reduced the dimensionality of this dataset by focusing on one question - ‘how many days in the last 2 weeks did you feel tired?’ with a threshold of ≥10/14 days using the median.

We chose a simplified approach with CUCQ32 for two reasons – to harmonize our analyses across the 3 IBD cohorts and to increase coverage for a large enough dataset for subsequent analyses. Here, we found 75.2% (866/1151), 43.6% (463/1061) and 14.2% (48/339) responses in active, remission and non-IBD controls respectively have ≥10/14 days fatigue days. The definition of fatigue by Markowitz et al. consisting of three components: the perception of generalized weakness, manifesting as inability or difficulty to initiate activities; quick fatigability and reduced capacity to maintain activities; and mental fatigue resulting in difficulty with concentration, emotional stability and memory; have been widely accepted as gold standard^14^. However, the simplified definition adopted in our study agrees with data from many other clinical studies on the prevalence of IBD fatigue^15, 16^.

Given the breadth of our dataset, we employed 6 different ML models that incorporated ∼100 different variables. Using different ML approaches to incorporate all available data in patient with active IBD and in remission; and as a combined group, we built in data fields such as seasonality, BMI, gender and drug therapy; and ML models generally achieve reasonable performance in predicting IBD-associated fatigue. However, this is more difficult in remission. This agrees with recent studies, there are no clear predictive factors of IBD-associated fatigue^17^ and point towards the enormous ‘hidden’ pathobiological component in this presentation. Many interventional studies have been stymied by the problem of patient heterogeneity – ‘are all patients tired in the same way?’^18^.

Our work has defined a starting point towards a clear data pathway and structure that with more data, may potentially identify different populations of fatigue phenotype where scientific studies can be designed around these specific patient groups. Extending from our point, a limitation of our study is the lack of molecular data such as genetics, microbiome and metabolic read-outs; and pertinent information such as tryptophan metabolites, diet and co-morbidities to complement our clinical predictive modelling. The acquisition of these molecular data is currently in progress in our laboratory. Having established our data pathway and structure as discussed above, we will build these lines of data in our ongoing work.

We have engendered a strong patient-involvement with participation of our patient groups in study design, conduct and analysis at all stages of our work. This is a creative juxtaposition of our patients and the adoption of machine-learning (as a step towards artificial intelligence methods) to advance of analysis of complex presentation such as fatigue in larger datasets. Recent studies have shown that routinely collected data in the clinic and in trial settings, can be leveraged by these methods to uncover hitherto new knowledge or function^19–22^. This is a new area with an immense pace of progress and a considered approach with close partnerships with patient groups is essential.

In conclusion, we show the significant impact of fatigue on wellbeing, even in remission. Our data provides credible support for the utility of patient reported outcomes as endpoints for future translational scientific research in this area. In addressing IBD-associated fatigue, we show our conceptual ability to reduce the dimensionality (thus flattening out the heterogeneity of IBD-associated fatigue) as the first-pass mechanism to identify the patients that are significantly affected by fatigue in active disease and in remission. This simplified approach will allow the future incorporation of multiple streams of complex scientific metadata (such as genetics and microbiome for example) for larger scale analyses at a cohort level. There are many better scientific tools to study central and peripheral fatigue – from imaging, metabolism and mitochondrial function. We envisage that our work provides a step towards identifying clear groups of IBD patients that will benefit from deeper human experimental studies to identify pathobiological mechanisms that can be therapeutically targeted and shifting away from a wholly symptom-based classification of IBD-wellbeing and fatigue.

## Methodology

### Research governance and ethical approvals

Investigation into Gastrointestinal Damage Associated Molecular Patterns (GI-DAMPs) is a cross-sectional IBD study with ethical approvals by East Scotland Ethics Committee REC 18/ES/0090 and R&D 2018/0197. Mitochondrial DAMPs as mechanistic biomarkers of mucosal inflammation in Crohn’s Disease (MUSIC study; www.musicstudy.uk) is a prospective IBD cohort study with ethical approvals by East Scotland Ethics Committee REC 19/ES/0087 and R&D No: 2019/0325. Both are mechanistic biomarker studies where patients provide specific consent and CUCQ32 data are routinely collected. All consented clinical and research data are anonymised and held in a secured database with no identifiable patient information.

### CUCQ32

The Crohn’s and Ulcerative Colitis Questionnaire-32 (CUCQ32) is a derivative work of the Inflammatory Bowel Disease Questionnaire (IBDQ), created and owned by McMaster University. CUCQ was used and developed by Swansea University for academic and non-commercial clinical purposes in the National Health Service (NHS). All other uses, modifications, and derivatives, including any translations and e-conversion for the CUCQ and its versions, require written permission from McMaster University. The average online completion time for CUCQ32 was 11 minutes.

### Clinical disease activity

Assessment of the patient’s current disease activity was performed using the Harvey– Bradshaw Index (HBI) for CD^23^ and Simple Clinical Colitis Activity Index (SCCAI) for UC^24^ (Appendix 2). The clinician conducting the study visit also recorded a global yes/no value for whether the patient had symptoms of active IBD (‘has_active_symptoms’ variable).

### Online CUCQ32 data

Using our patient and public networks as earlier described, all online data collected are wholly anonymous with no identifiable information. The categorization of disease activity, IBD (UC, CD, IBD-U) and non-IBD status are self-reported with no recourse for the research group to verify independently. For participants of our online survey, there is a further option to provide specific comments. Initial screening was performed by RH and GTH for identifiable information. All data is held in a secured database held in the Edinburgh IBD Science group, University of Edinburgh computer server.

### Machine learning methods

The machine learning pipeline used is summarised in Figure 4a. The required dependencies, anonymised dataset and code required to reproduce the above pipeline is available for inspection at our GitHub repository (<u>github.com/1-</u> <u>gut/machine_learning_for_ibd_fatigue</u>).

Python 3.11.9 was the base runtime used. Machine learning was undertaken using scikit-learn v1.5.2, scipy 1.14.1, xgboost 2.1.2, pandas 2.2.3, numpy 2.0.2, shap 0.46.0. Data was first pre-processed. Missing data was imputed with a variety of strategies depending on the nature of the variable, for example, non-normally distributed continuous variables such as crp, albumin, calprotectin, were imputed with the median. Subsequently, categorical features were converted into binary columns via one-hot-encoding. Numerical features were scaled with scikit-learn’s StandardScaler (which standardizes the mean to 0 and preserves the variance). Feature engineering was performed to calculate season variable (based on date of CUCQ32 questionnaire) and diagnosis duration in weeks. The dataset was then split 70:30 into training and testing sets. The training set was then used to generate 5-fold cross validation datasets using a GroupKFold strategy ensuring that the same study participant does not appear across training and validation datasets.

Six different machine learning models were used - XGBoost, random forest classifier, AdaBoost, multilayer perceptron classifier, support vector classifer and logistic regression. A brief description of each model is provided in Table 4. Hyperparameter tuning was performed on all six models to search for the best model hyperparameters. The best hyperparameters were then used to create the final machine learning models. These models were then evaluated on the testing set which was set apart at the beginning. AUC curves were generated to analyse the predictive capability of the models on the testing set.

**Table 4:** Summary of machine learning algorithms used.

| Algorithm | Description |
| --- | --- |
| Logistic regression | Takes input variables, calculates a weighted sum, converts it into probability using a sigmoid function |
| XGBoost | Builds a series of decision trees sequentially, with each new tree correcting errors made by the previous one, using a method known as gradient boosting |
| AdaBoost | Trains a series of weak classifiers, with each one focused on correcting the mistakes of the previous one and the final model is a combination of these classifiers |
| Random Forest | Creates a 'forest' of decision trees, each trained on a random subset of data. Final prediction is made by averaging the predictions of all the trees. |
| Support Vector Classifier | Finds the hyperplane in n-dimensional space that best separates the different classes of data. The hyperplane is chosen to maximize the margin between the classes |
| Multilayer Perceptron Classifier | A type of artificial neural network that has multiple layers of neurons – an input layer, one or more hidden layers and an output layer. |

### Data and code availability

Anonymised data and code are available at https://github.com/1-gut/machine_learning_for_ibd_fatigue.

### Patient-clinical researcher model

Our patient group is derived from Edinburgh IBD Science patient volunteer group (led by MH; including AT, PK, EP, and DD) that has been active since 2019. PK and EP are the UK charitable organization Crohn’s Colitis UK patient research champions (https://crohnsandcolitis.org.uk/). Under the oversight of RH and GTH (IBD clinicians), our IBD group has access to the online database of anonymized IBD patients’ inputs. Throughout the duration of our study, our patient-clinician group meet regularly to discuss study design, data analysis, presentation and direction. Our patient group has written the lay summary for this study and generated a patient-led paper into the IBD wellbeing (REF) under the Sponsor of GTH as the principal investigator and the guarantor.

### Statistical analysis

Basic descriptive statistics and statistical analyses were performed using a combination of GraphPad Prism 10.4.0, R version 4.4.1 and Python 3.11.9/SciPy 1.14.1. T-tests were used to compare normally distributed continuous variables. Chisquare tests were used to compare categorical variables. One-way ANOVA was performed for comparisons of continuous variables across three or more groups, with post-hoc pairwise comparison testing utilizing the Bonferroni correction. Pairwise Spearman correlations of CUCQ32 Question 5 and all other CUCQ32 questions was performed using R, with p-values Benjamini-Hochberg–adjusted.

## Acknowledgement

This work is part of the MUSIC IBD study funded by The Leona M. and Harry B. Helmsley Charitable Trust (G-1911-03343) and Anne Ferguson Memorial Fund to GTH.

## Supplementary Figures

**Supplementary Figure 1a.**
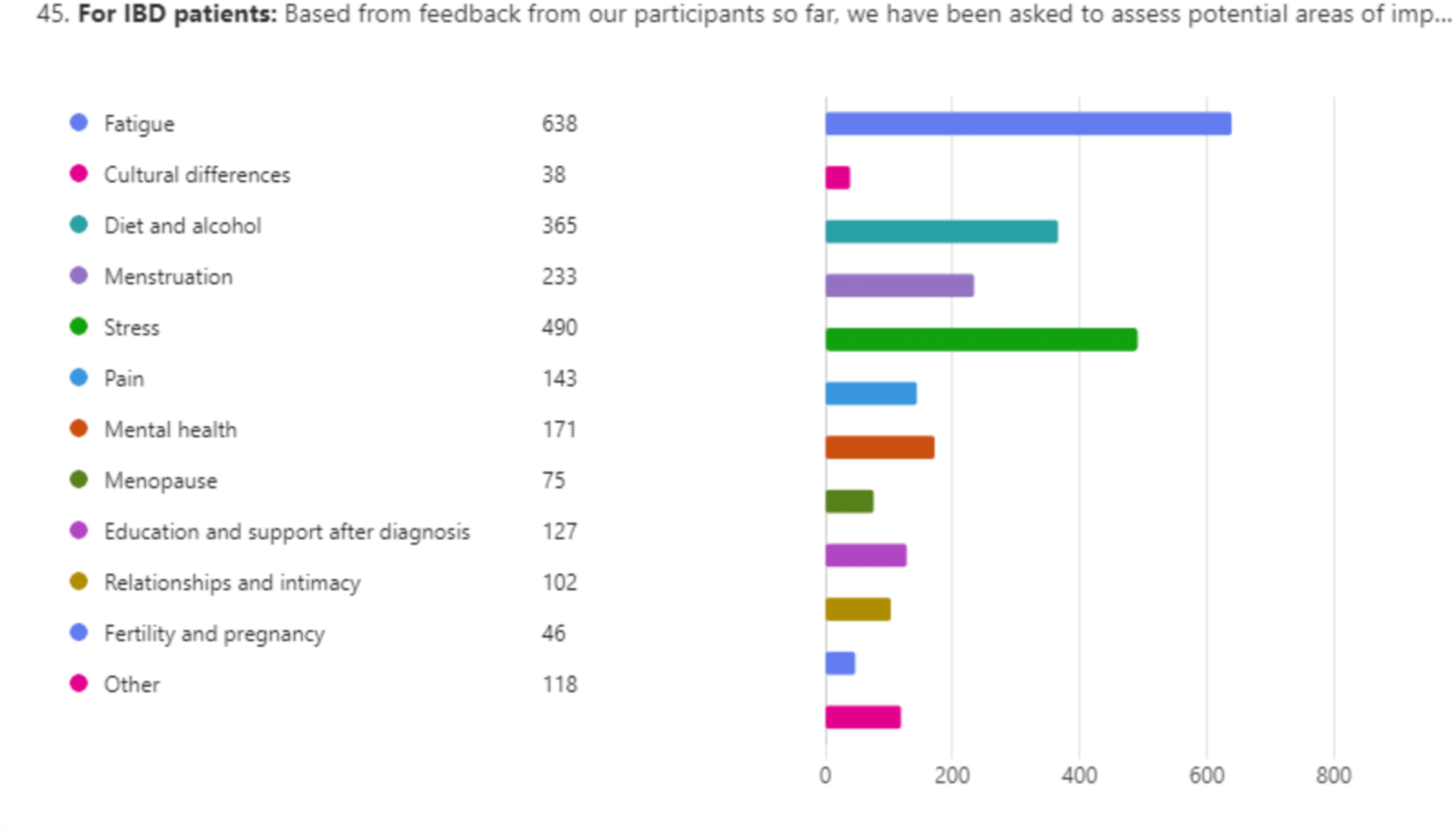
Feedback from online survey about areas participants would like prioritisation for further research

**Supplementary Figure 2a.**
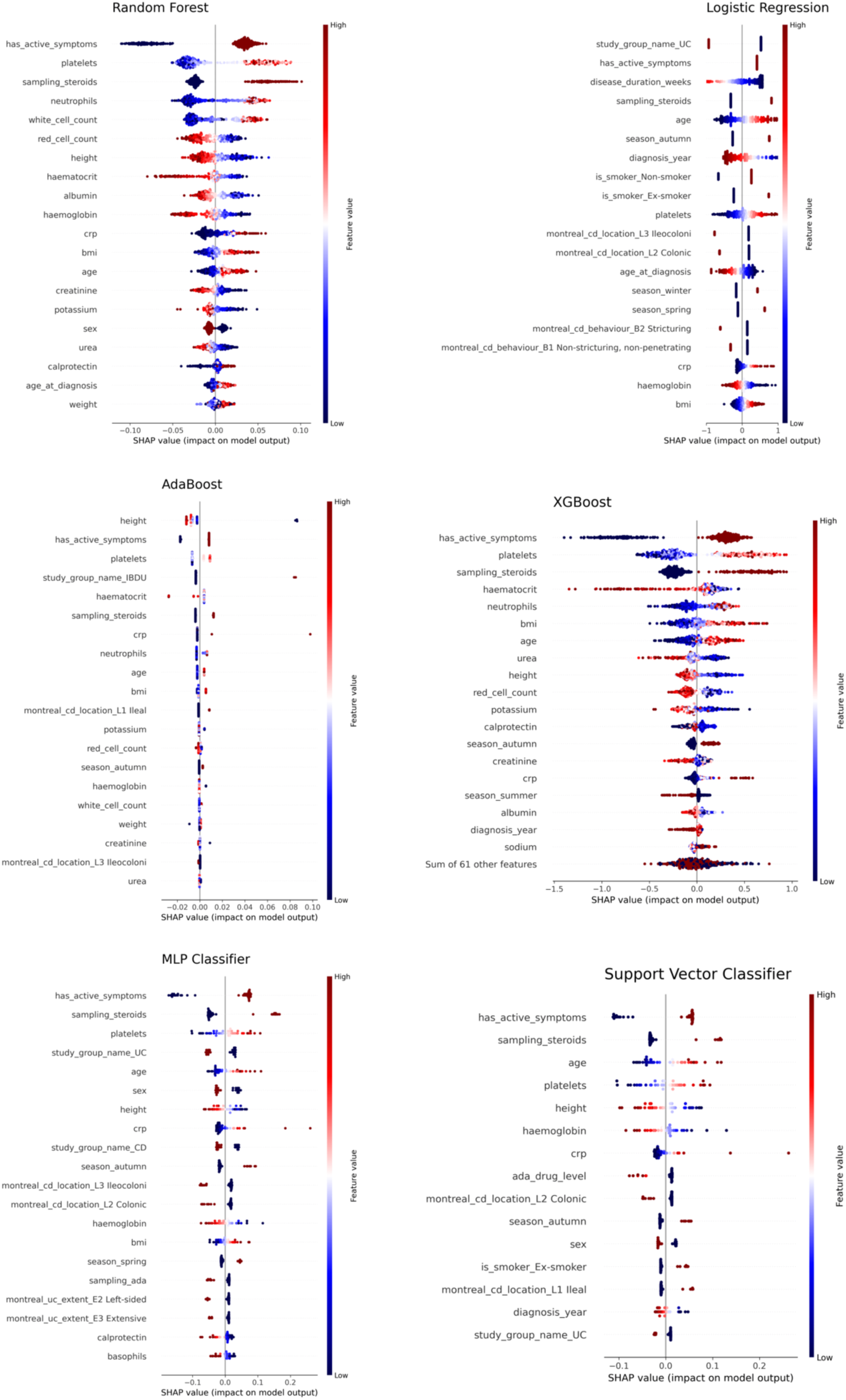
SHAP analysis for all models in the full IBD cohort

**Supplementary Figure 2b.**
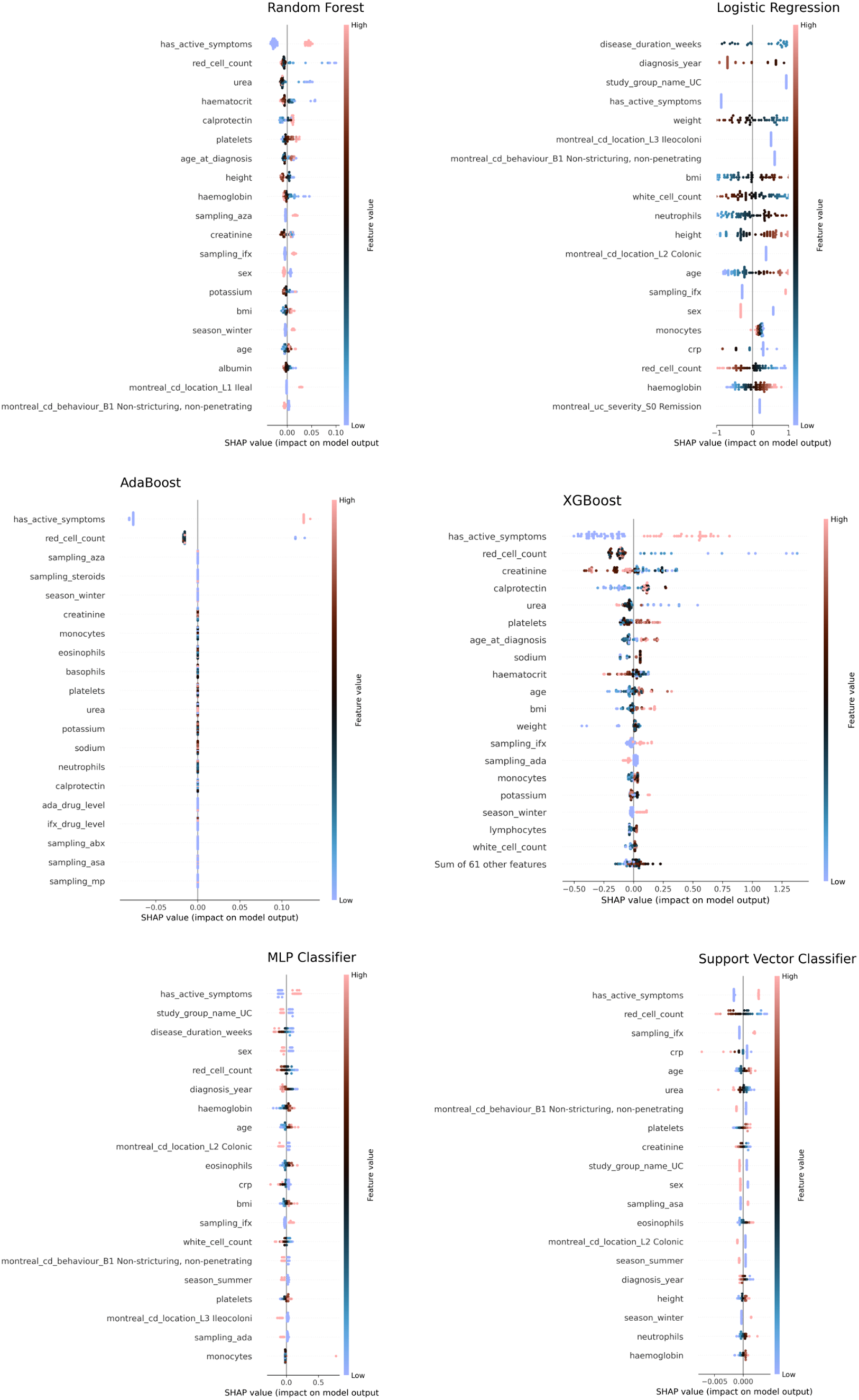
SHAP analysis for all models in the biochemical remission cohort

## Appendix

**Appendix 1:**
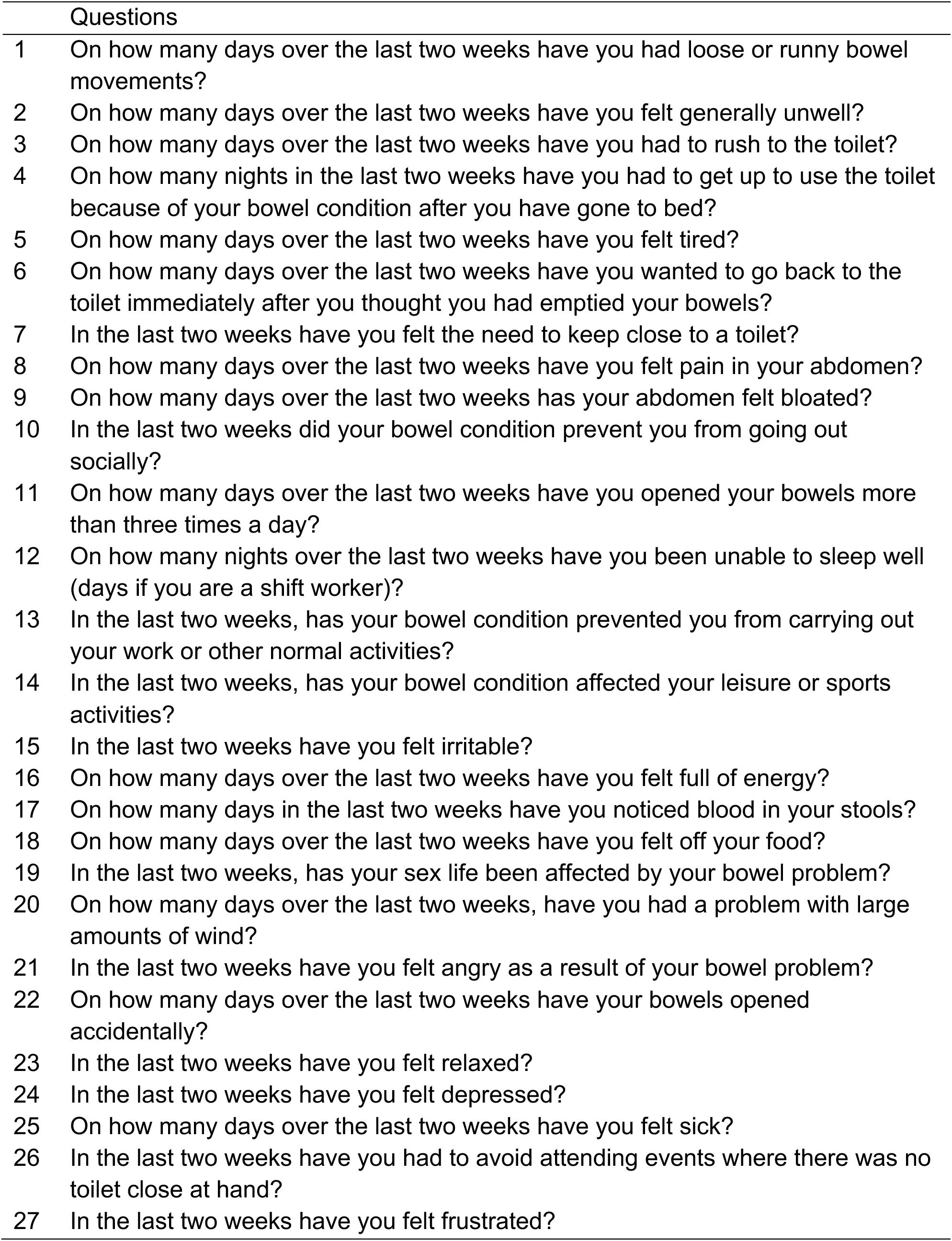
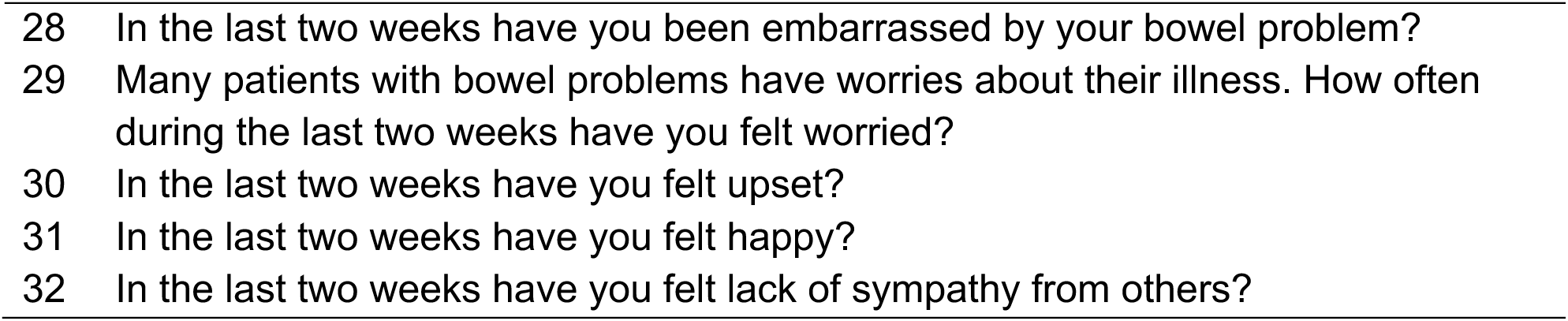
The CUCQ32 questions.

**Appendix 2:** Harvey Bradshaw Index (HBI) and Simple Crohn’s and Colitis Activity Index (SCCAI)

Harvey-Bradshaw Index (Crohn's)
|  | 0 | 1 | 2 | 3 | 4 |
| --- | --- | --- | --- | --- | --- |
| **General wellbeing** | Very well | Slightly below par | Poor | Very poor | Terrible |
| **Abdominal pain** | None | Mild | Moderate | Severe |  |
| **Abdominal mass** | None | Dubious | Definite | Definite and tender |  |
| **Arthralgia** | No | Yes |  |  |  |
| **Uveitis** | No | Yes |  |  |  |
| **Erythema nodosum** | No | Yes |  |  |  |
| **Aphthous ulcers** | No | Yes |  |  |  |
| **Pyoderma gangrenosum** | No | Yes |  |  |  |
| **Anal fissures** | No | Yes |  |  |  |
| **New fistula** | No | Yes |  |  |  |
| **Abscess** | No | Yes |  |  |  |

Simple Clinical Colitis Activity Index (UC)
|  | 0 | 1 | 2 | 3 | 4 |
| --- | --- | --- | --- | --- | --- |
| **General wellbeing** | Very well | Slightly below par | Poor | Very poor | Terrible |
| **Bowel frequency (day)** | 1-3 | 4-6 | 7-9 | >9 |  |
| **Bowel frequency (night)** | 0 | 1-3 | 4-6 |  |  |
| **Urgency of defecation** | None | Hurry | Immediately | Incontinence |  |
| **Blood in stool** | None | Trace | Occasionally frank | Usually frank |  |
| **Arthralgia** | No | Yes |  |  |  |
| **Uveitis** | No | Yes |  |  |  |
| **Erythema nodosum** | No | Yes |  |  |  |
| **Pyoderma gangrenosum** | No | Yes |  |  |  |

**Total = \_\_\_\_\_ (0-19)** (≤2 remission)

**Appendix 3:** Paired correlation analyses of Question no 5, (‘How many days in the last 2 weeks did you feel tired?’) against 31 CUCQ32 questions. Spearman-rank analyses, Benjamini-Hochberg corrected.

| CUCQ32 Question Number | How many days are you tired in the last 2 weeks (Q5)? |
| --- | --- |
| 1 | 1.33E-69 |
| 2 | 3.19E-147 |
| 3 | 2.01E-85 |
| 4 | 1.40E-51 |
| 6 | 3.90E-67 |
| 7 | 1.96E-82 |
| 8 | 2.44E-102 |
| 9 | 7.58E-93 |
| 10 | 3.85E-86 |
| 11 | 1.16E-65 |
| 12 | 4.80E-103 |
| 13 | 6.59E-79 |
| 14 | 3.45E-90 |
| 15 | 2.03E-82 |
| 16 | 3.25E-142 |
| 17 | 1.89E-35 |
| 18 | 8.76E-67 |
| 19 | 1.23E-63 |
| 20 | 1.39E-46 |
| 21 | 7.60E-47 |
| 22 | 3.08E-21 |
| 23 | 3.07E-49 |
| 24 | 1.91E-48 |
| 25 | 5.80E-57 |
| 26 | 1.47E-47 |
| 27 | 4.80E-73 |
| 28 | 1.26E-53 |
| 29 | 1.73E-57 |
| 30 | 8.37E-63 |
| 31 | 1.61E-28 |
| 32 | 2.97E-09 |

**Appendix 4:** List of input features for machine learning after data transformation.

| Clinical | Laboratory | Drug Therapy |
| --- | --- | --- |
| Age | Haemoglobin | Steroids |
| Sex | Red cell count | Antibiotics |
| Height | White cell count | Aminosalicylates |
| Weight | Neutrophils |  |
| BMI | Lymphocytes | Azathioprine |
|  | Monocytes | Mercaptopurine |
| Age at diagnosis | Eosinophils | Methotrexate |
| Date of diagnosis | Basophils | Ciclosporin |
| Disease duration | Platelets |  |
| Diagnosis group |  | Infliximab |
| Montreal classification | Urea | Adalimumab |
| Previous extra-intestinal manifestations | Creatinine | Vedolizumab |
|  | Sodium | Ustekinumab |
|  | Potassium | Risankizumab |
| Has active symptoms |  |  |
| Smoking Status | Albumin | Tofacitinib |
| Season of CUCQ completion | C-reactive protein | Filgotinib |
|  | Stool calprotectin | Upadacitinib |
|  | Infliximab antibody status |  |
|  | Infliximab drug level |  |
|  | Adalimumab antibody status |  |
|  | Adalimumab drug level |  |

## Notes

### Competing Interest Statement

The authors have declared no competing interest.

### Clinical Protocols

https://clinicaltrials.gov/study/NCT04760964

### Author Declarations

Investigation into Gastrointestinal Damage Associated Molecular Patterns (GI-DAMPs) is a cross-sectional IBD study with ethical approvals by East Scotland Ethics Committee REC 18/ES/0090 and R&D 2018/0197. Mitochondrial DAMPs as mechanistic biomarkers of mucosal inflammation in Crohn's Disease (MUSIC study; www.musicstudy.uk) is a prospective IBD cohort study with ethical approvals by East Scotland Ethics Committee REC 19/ES/0087 and R&D No: 2019/0325.

